# Impact of race, sex and age on the probability of pancreatic cancer among patients with newly diagnosed diabetes

**DOI:** 10.1101/2024.12.13.24319002

**Authors:** Elham Afghani, Bryan Lau, Laura Becker, Michael G Goggins, Alison P. Klein

**Affiliations:** Division of Gastroenterology, Department of Medicine, The Johns Hopkins Medical Institutions, Baltimore, Maryland, USA; Department of Epidemiology, Johns Hopkins University Bloomberg School of Public Health, Baltimore, MD 21205, USA; Division of Internal Medicine, Department of Medicine, The Johns Hopkins School of Medicine, Baltimore, Maryland, USA; Optum Labs, Eden Prairie, MN; Department of Oncology, Sidney Kimmel Comprehensive Cancer Center at Johns Hopkins, Baltimore, MD 21231, USA; Department of Pathology, Sol Goldman Pancreatic Cancer Research Center, Department of Pathology, The Johns Hopkins School of Medicine, Baltimore, Maryland, USA

**Keywords:** Pancreatic adenocarcinoma, diabetes, race, risk

## Abstract

**Background:** Pancreatic cancer diagnoses are frequently preceded by a new diabetes diagnosis. Screening individuals newly diagnosed with diabetes could enable earlier pancreatic cancer detection. We sought to estimate the risk of pancreatic cancer by age, sex, race and time since diabetes diagnosis.

**Methods:** Johns Hopkins Medicine conducted this de-identified claims-based cohort study using the Optum Labs Data Warehouse (OLDW). Enrollees from 1/2008–9/2018 were identified as non-diabetic or newly diagnosed diabetics and time to pancreatic cancer analysis was conducted using a flexible Weibull model. Diabetes and cancer were defined using ICD-9/10 codes.

**Results:** Our risk set included 4,732,313 individuals (424,129 newly diabetic) in 5,844,934 enrollment periods. Individuals with newly diagnosed diabetes were at an increased hazard ratio (HR) of pancreatic cancer but this effect waned over time. The HR of pancreatic cancer following a diabetes diagnosis was higher in younger individuals and varied by race (lower HR in non-White individuals) (p<0.01, main effects and interactions). Thus, the probability of pancreatic cancer following a diabetes diagnosis was dependent on age, race, and sex. For example: the 1-year probability of pancreatic cancer in a white male aged 75 was 0.45% (95%CI 0.41%-0.49%) if they were newly diagnosed with diabetes and 0.090% (95%CI 0.084%-0.096%) fi they were free of diabetes. In contrast, risk was lower if they were age 55 at 0.15% (new-diabetic, 95%CI 0.13%-0.16) and 0.022% (diabetes free, 95%CI 0.020%-0.023%). The HR of pancreatic cancer for individuals with newly diagnosed diabetes compared to those free of diabetes was highest 1 month after diagnosis (HR=9.6 and 14.7 for a 75 and 55 year old while male, respectively) but decreased in the following months, with a ∼39% reduction in HR from 1- to-3 months, ∼17% from 3 -to-6 months, and ∼14% from 6 month-to -1 year (p<0.01).

**Conclusions:** Consideration of the age-race-sex specific probability of pancreatic cancer and time since diabetes diagnosis is necessary to when evaluating the risk of pancreatic cancer following a diabetes diagnosis.

## INTRODUCTION

Pancreatic cancer is the 3rd leading cause of cancer-related death in the United States with an estimated 60,430 new cases and 48,220 deaths in 2021^1^.Globally, the number of pancreatic cancer cases diagnosed annually has doubled since 1990, and age-adjusted incidence rates have increased from 5 per 100,000 person-years in 1990 to 5.7 per 100,000 person-years in 2017.^3^ Pancreatic cancer is usually clinically silent in its early stages. The advanced stage at diagnosis is a major factor in the low 5-year survival rates, currently 10%, the lowest of any major tumor site.^1, 2^

Studies have consistently identified an association between pancreatic cancer and diabetes.^4–6^ Long-standing diabetes is associated with an ∼2 fold increased risk of pancreatic cancer.^16^ However, many pancreatic cancer patients present with a recent diagnosis of diabetes^7–18^ and up to 85% of pancreatic cancer patients have diabetes or impaired fasting glucose.^9^ Resection of the tumor frequently leads to resolution of the diabetes, suggesting that this is a manifestation of the cancer^9^ and it may be one of the few early manifestations of an otherwise silent disease.

One of the first studies to examine pancreatic cancer risk in individuals with newly diagnosed diabetes was a retrospective study within the Mayo Clinic population which indicated up to 1% of patients develop pancreatic cancer within 3 years of their diabetes diagnosis.^13^ However, subsequent studies conducted with the VA system found the risk of pancreatic cancer to be lower, with <0.3% developing pancreatic cancer within 3 years of their diabetes diagnosis compared to 0.11% in patients without diabetes.^10, 11^ A recent study, using electronic health record (EHR) data within the Kaiser Permanente Southern California system, found that the risk of pancreatic cancer was increased 7-fold in those with diabetes diagnosed within one year of cancer diagnosis.^15^ Other studies have suggested a ∼4.5 to 5 increased odds of having pancreatic cancer in the year following a diabetes diagnosis.^14, 19^ However, these studies had a limited ability to examined how age, a major predictor of pancreatic cancer risk as well as race and sex impact the probability of a pancreatic cancer diagnosis subsequent to a diabetes diagnosis. A better understanding of the time dependent probability of a pancreatic cancer diagnosis following a diabetes diagnosis, and how age and race modifies this probability, is necessary to understand the understand the potential benefit of focused pancreatic cancer screening in individuals with a new diabetes diagnosis. This includes an understanding of the lead-time between a diabetes claim and subsequent cancer diagnosis reflecting the window of opportunity for earlier detection of pancreatic cancer.

There are ongoing efforts to enroll individuals with new diabetes diagnosis to determine how to effectively identify the subset with underlying pancreatic cancer.^20, 21^ A better understanding of how the probability of a pancreatic cancer diagnosis following a new diabetes diagnosis varies with age, race sex and time since diabetes diagnosis can provide insight into the potential role for earlier detection in this population. Furthermore, estimation of these risks using real-world data can provide insights beyond what can be learned in more controlled observational settings. The goal of this study was to use claims data to quantify the probability of a pancreatic cancer diagnosis as a function of time following the initial diabetes claim while accounting for age, race and sex to demonstrate the window for early detection. This informs the potential yield of pancreatic cancer screening interventions in the real-world settings of US individuals with health insurance.

## METHODS

Our claims based cohort was defined using de-identified administrative claims data from the Optum Labs Data Warehouse (OLDW). Since this study involved analysis of pre-existing, de-identified data, it was exempt from Institutional Review Board review. The database contains longitudinal health information on enrollees and patients, representing a mixture of ages, and geographies across the United States. The claims data in OLDW includes medical and pharmacy claims, and enrollment records for commercial and Medicare Advantage enrollees.^22^ Demographic information, health plan enrollment status, inpatient and outpatient medical encounters coded using the *International Classification of Diseases, Ninth Revision, Clinical Modification (ICD-9-CM),* the *International Statistical Classification of Diseases, Tenth Revision, Clinical Modification (ICD-10-CM),and Current Procedural Terminology, fourth edition (CPT-4)* and filled prescriptions (including the National Drug Code numbers, quantity dispensed, and days’ supply) were recorded for each patient (See STable 1 for codes used). Ethnicity is assigned by an external vendor who uses a rule-based system that combines analysis of first names, middle names, surnames, and surname prefixes and suffixes with geographic criteria. Optum Labs then assigns these ethnicity values into one of five compliance-determined race code values: W (Non-Hispanic White), B (Non-Hispanic Black), H (Hispanic), A (Asian), and U (Unknown).

Our cohort was limited to individuals aged 40-88 years with a minimum 12 months of continuous enrollment (with both medical and pharmacy benefits) from 2007 to 2018. Using claims data we identified two groups individuals 1) those with an incident diabetes diagnosis and 2) individuals without diabetes claims. *Individuals with newly diagnosed diabetes were defined as individuals with*: 1) two non-diagnostic medical claims for diabetes at least 30 days apart, or 2) one non-diagnostic medical claim for diabetes and a prescription filled for an oral anti-diabetic or insulin medication; the earliest claim associated with diabetes was identified as the index date. ICD codes covering all subtypes of diabetes, and pharmaceutical claims for hypoglycemic medications and insulin were considered evidence of diabetes (see STable 1 for codes used). To exclude individuals with prevalent diabetes, we required minimum 1-year period without evidence of diabetes (none of the diabetes defining claims) prior to the index data. *Non-diabetic comparison group* was defined as individuals without evidence of diabetes claims (no non-diagnostic medical claims nor diabetic prescriptions). For these individuals, if they had no evidence of diabetes at any time before the index date, (defined as enrollment start date plus a randomly generated number of days based upon a Gamma distribution (alpha=1.0, beta=1100) was used at the start time in time to event analysis to mirror the distribution in the incident diabetes cohort). Individuals may contribute more than one enrollment period (i.e. individuals may enter and exit the medical plan, each time continuous enrollment starts, a wash-out period of 1-year was evaluated for study inclusion/exclusion criteria) as such non-diabetic individual may later become diabetic and thus contribute person-time accordingly. For both groups, follow-up time was censored at the first non-diagnostic claim for pancreatic cancer based on ICD-9/10 codes (STable 1), end of health plan enrollment or 6 years after index date. (See STable 2 for details). Additional exclusions include pregnancy in the first year of baseline period (due to the possibility of gestational diabetes), diagnosis of cancer in the year prior to the index date, and those diagnosed with pancreatic neuroendocrine tumor.

## STATISTICAL ANALYSIS

Time-to-event analysis was conducted using a parametric spline-based survival model which allowed for a better fit to the data over a traditional Cox survival model. Flexible parametric spline models with 1-4 knots^23^ were fit to the data; covariates were plotted individually to verify fit, and multivariable model fit was determined using the AIC (Akaike Information Criterion). The best-fit model had 2 internal knots and 2 ancillary spline parameters modeled as linear functions of time (diabetes diagnosis or start of follow-up) and age, where the effects of diabetes and age are arbitrarily flexible functions of time, with interaction effects of diabetes on age and race. Hazard rates and survival estimates derived from the multivariable model were used to estimate race, sex, and age specific hazard ratio (*h*_diabetes_/*h*_no-diabetes_), and probability estimates ((1-*S(t)*)*100) where *S(t)* is the survival function estimate at time *t* for either diabetics or non-diabetics as appropriate at specified time points.

Proportional hazards assumptions and Kaplan-Meier curves were examined in RStudio^24^ using the survival package.^25^ Time-to-event analyses were conducted in R using the flexsurv and flexsurvspline packages.^26^ Plots were produced in R using the ggplot2 package.^27^

## RESULTS

Of 5,844,934 enrollment periods across 4,732,313 individuals identified in the study population, there were 424,129 enrollment periods after a new diabetes diagnosis (7.3%) and 5,420,805 periods for non-diabetic individuals. Table 1 shows the characteristics of the enrollment periods. The mean age of enrollment periods was 55 (range 40-88) years, with an older mean age for the enrollment periods after a new diabetes diagnosis (59; range 40-88), than enrollment periods among individuals without diabetes (54, range 40-88 years). The median length an enrollment period was 1.7 years (range 0-6 years) excluding wash-out year. In total, 5,713 individuals were diagnosed with pancreatic cancer during follow-up.

**Table 1:**
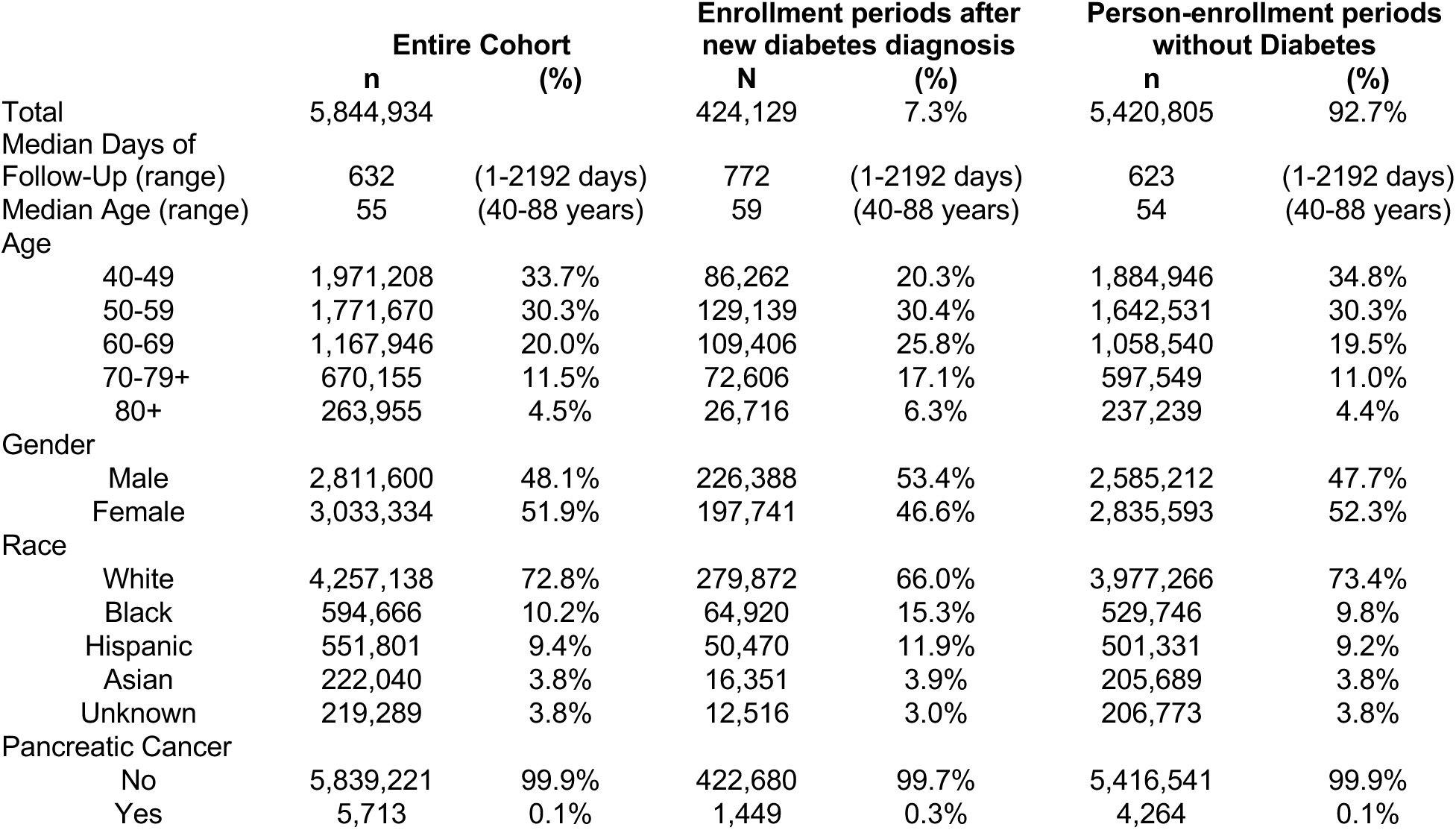
Cohort Demographics.

We sought to determine how age, race, sex and time since diabetes diagnosis impacted the probability of a pancreatic cancer diagnosis over time. To estimate time to pancreatic cancer diagnosis in relation to age, sex and race, we included each of these as predictors along with their interactions in flexible Weibull survival model (Hazard Ratios presented in Table 2 and Model parameters in Table 3). The time-scale was days following index date, which for incident diabetics reflects the date first diabetes claim. The hazard of pancreatic cancer in individuals remaining diabetes free was relatively constant over the study period (Table 3) and was lower than individuals with diabetes. Among individuals without diabetes the hazard ratio for pancreatic cancer was higher (HR 1.29 (95%CI 1.22-1.36) in males compared to females and was higher in Black individuals compared to White individuals (HR=1.27 (95%CI 1.15-1.39). Whereas, hazard ratios were similar in Asian, Hispanic and White individuals (HR 0.92 (95%CI 0.76-1.10) and HR=1.02 (95%CI 0.91-1.15) for Asian vs White and Hispanic vs White, respectively. In addition, the hazard rate of pancreatic cancer in individuals without diabetes increased with age but was not constant over all ages (see Table 3 for model estimates). In contrast, the hazard ratio of pancreatic cancer in individuals with newly diagnosed diabetes compared those remaining diabetes free was highest immediately following the diabetes diagnosis and decreased as time following diabetes diagnosis increased (see Table 3 and Figure 1 for more estimates). On average the hazard ratio comparing individuals with new diabetes compared to those remaining diabetes free decreased ∼39% between 1 month and 3 months, ∼17% from 3 months to 6 months and ∼14% from 6 months to 1 year. In addition, the hazard of pancreatic cancer due to a diabetes diagnosis varied by age and race (Figure 1, Table 2, Table 3). The hazard ratio of pancreatic cancer due to diabetes decreased with age (Figure 1, Table 2), and was highest in White individuals compared with Black, Asian or Hispanic individuals.

**Figure 1:**
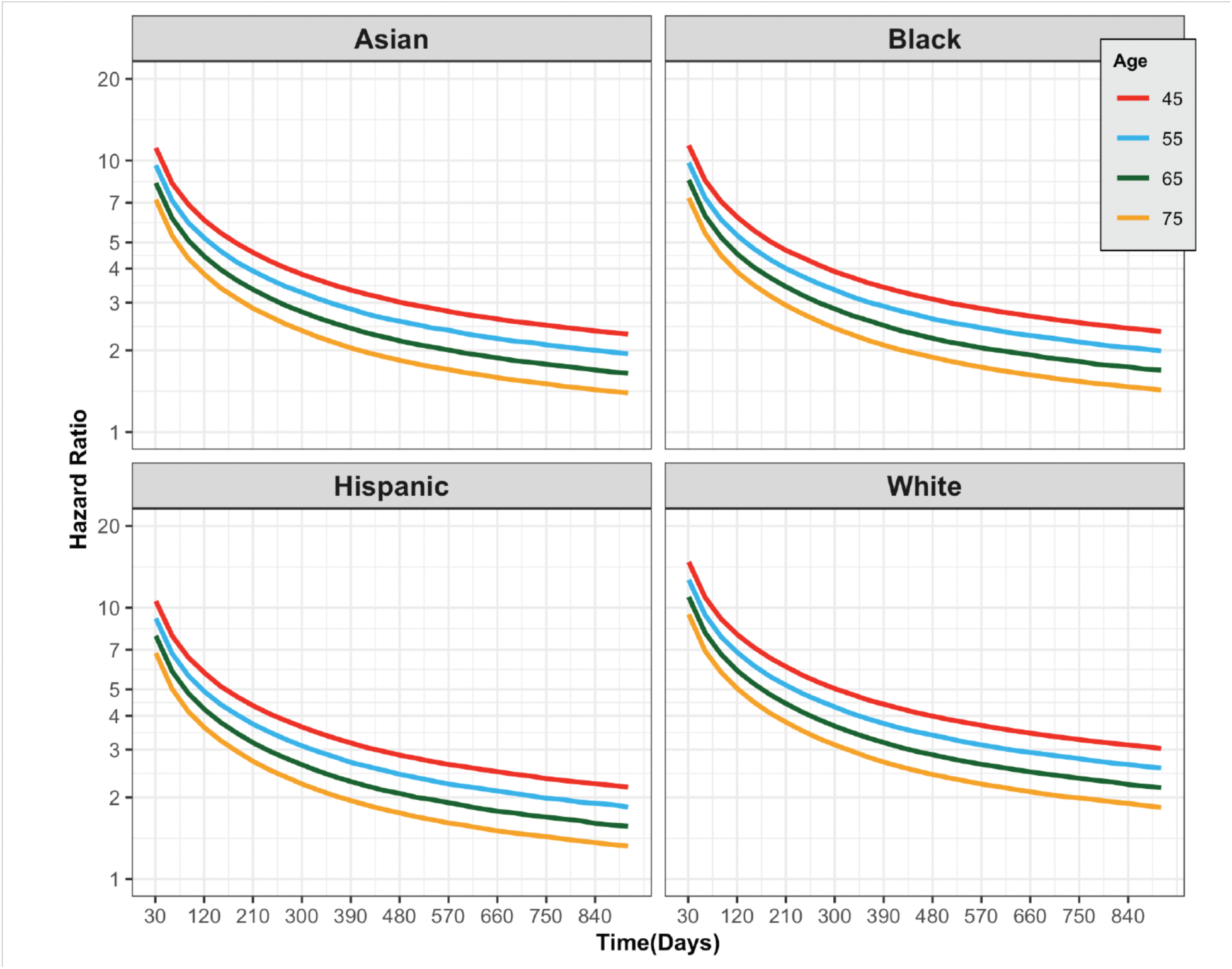
Hazard ratio of pancreatic cancer in individuals with new diabetes vs. non-diabetics by age, race and days since diabetes diagnosis/cohort entry

**Table 2:**
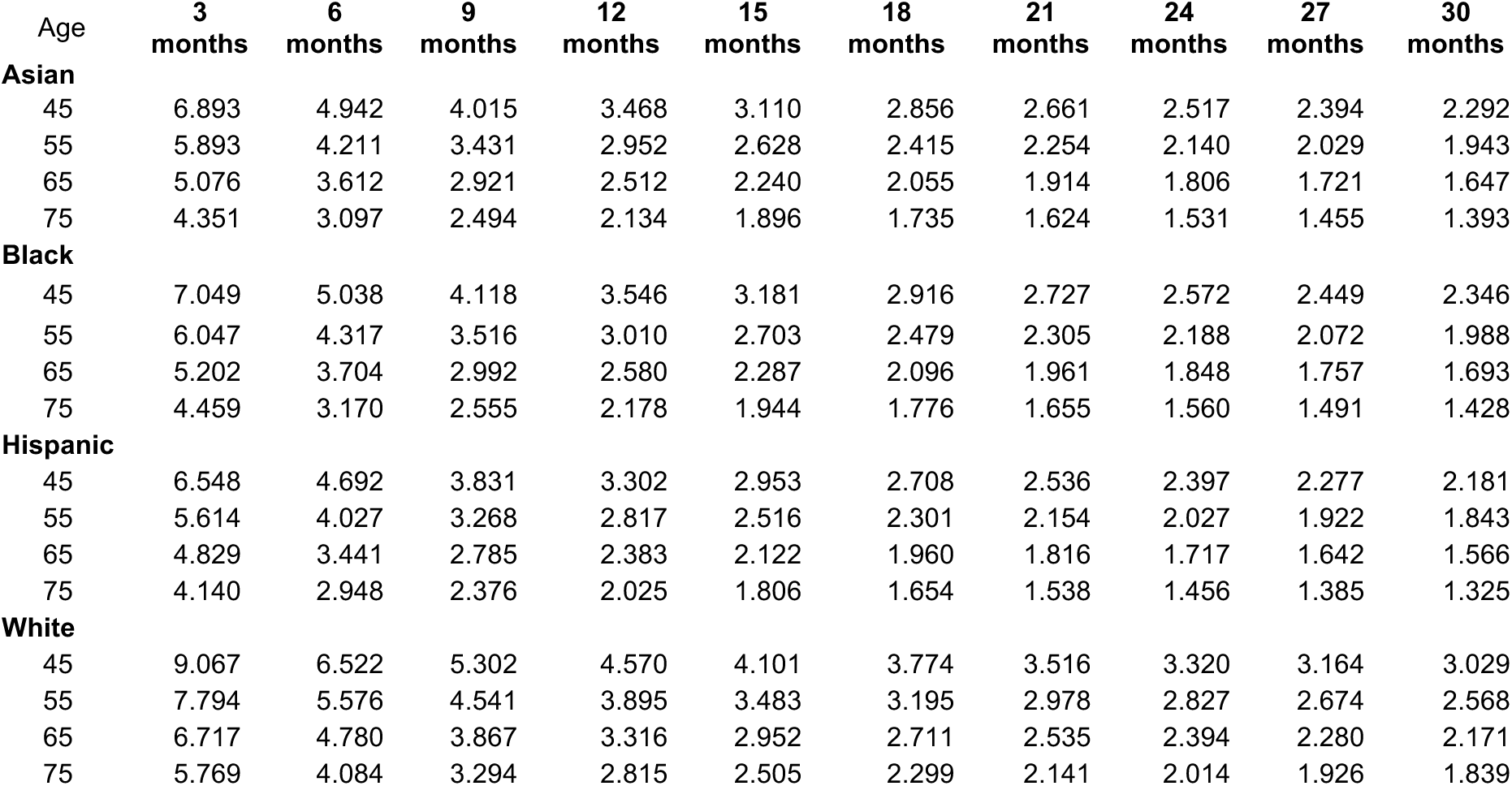
Hazard ratio for Pancreatic Cancer in individuals with newly diagnosed diabetes compared to non-diabetics by age, race and days since diabetes diagnosis/cohort entry.

**Table 3:**
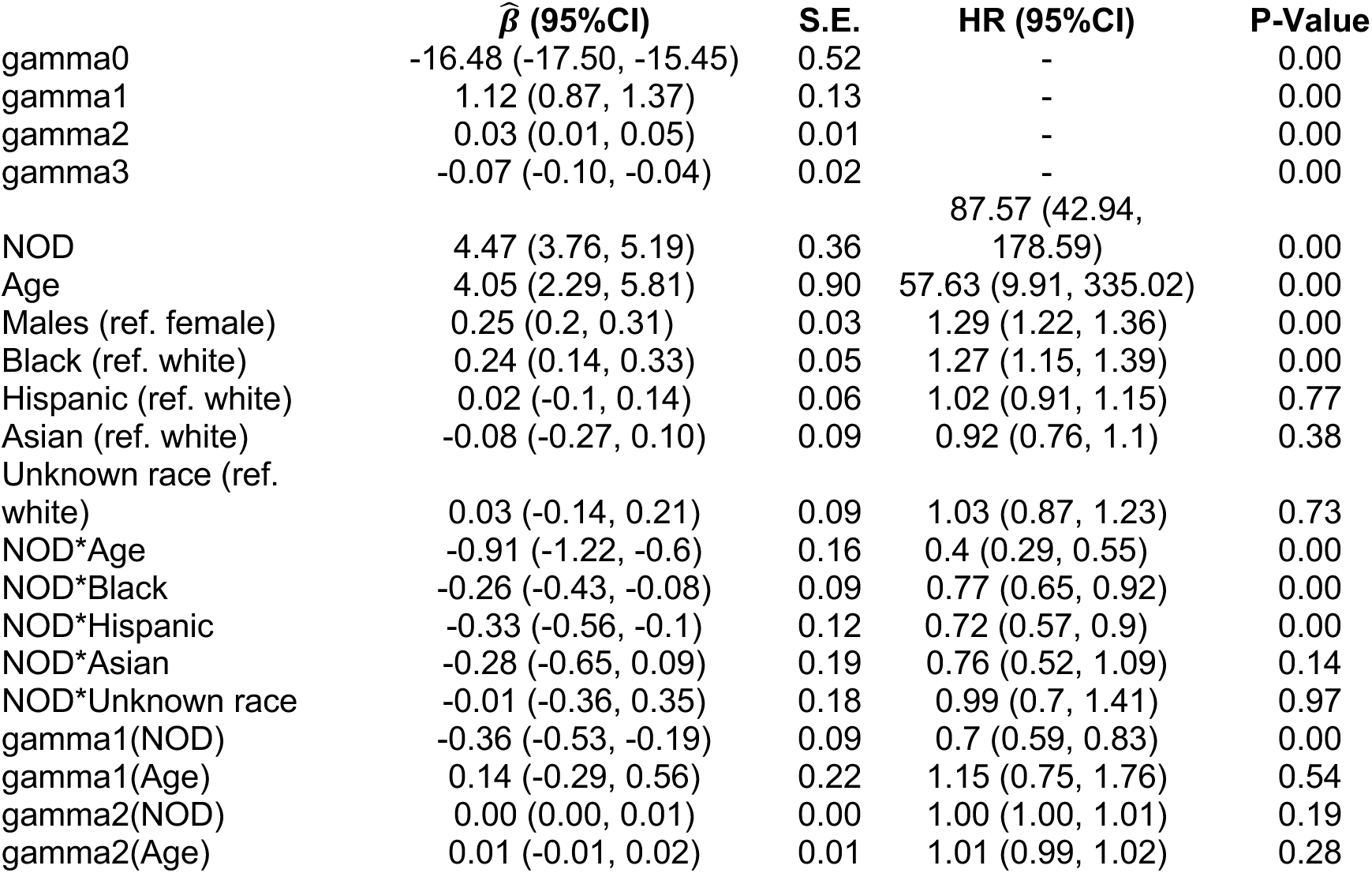
Flexible survival spline hazards model parameter estimates.

Figure 2A and 2B shows the predicted cumulative probability of pancreatic cancer by time since selection/diabetes diagnosis. The curves represent specific ages by diabetes status and panels by race. For clarity, cumulative probability estimates at 3 months, 6 months, 12 months, 24 months and 36 months timepoints are also shown in Table 4. The cumulative probability was highest in older individuals with a new diabetes diagnosis, with a cumulative probability of 0.450% (95%CI 0.413-0.489) at one year after diabetes diagnosis in a 75-year old white male. While the hazard ratio of pancreatic cancer in individuals with diabetes was highest in White individuals. However, cumulative pancreatic cancer probability in Black individuals with a new diabetes diagnosis was generally comparable to White individuals with a new diabetes diagnosis at all timepoints, with a cumulative probability of 0.422% (95%CI 0.367-0.485) at one years after diabetes diagnosis in a 75-year old black male. In individuals remaining diabetes free, the cumulative probability of pancreatic cancer was higher in Black individuals compared with White individuals cumulative probability in one year 0.112% (0.102, 0.124) and 0.090% (0.084, 0.096) in a 75 year old non-diabetic black and white male respectively.

**Figure 2:**
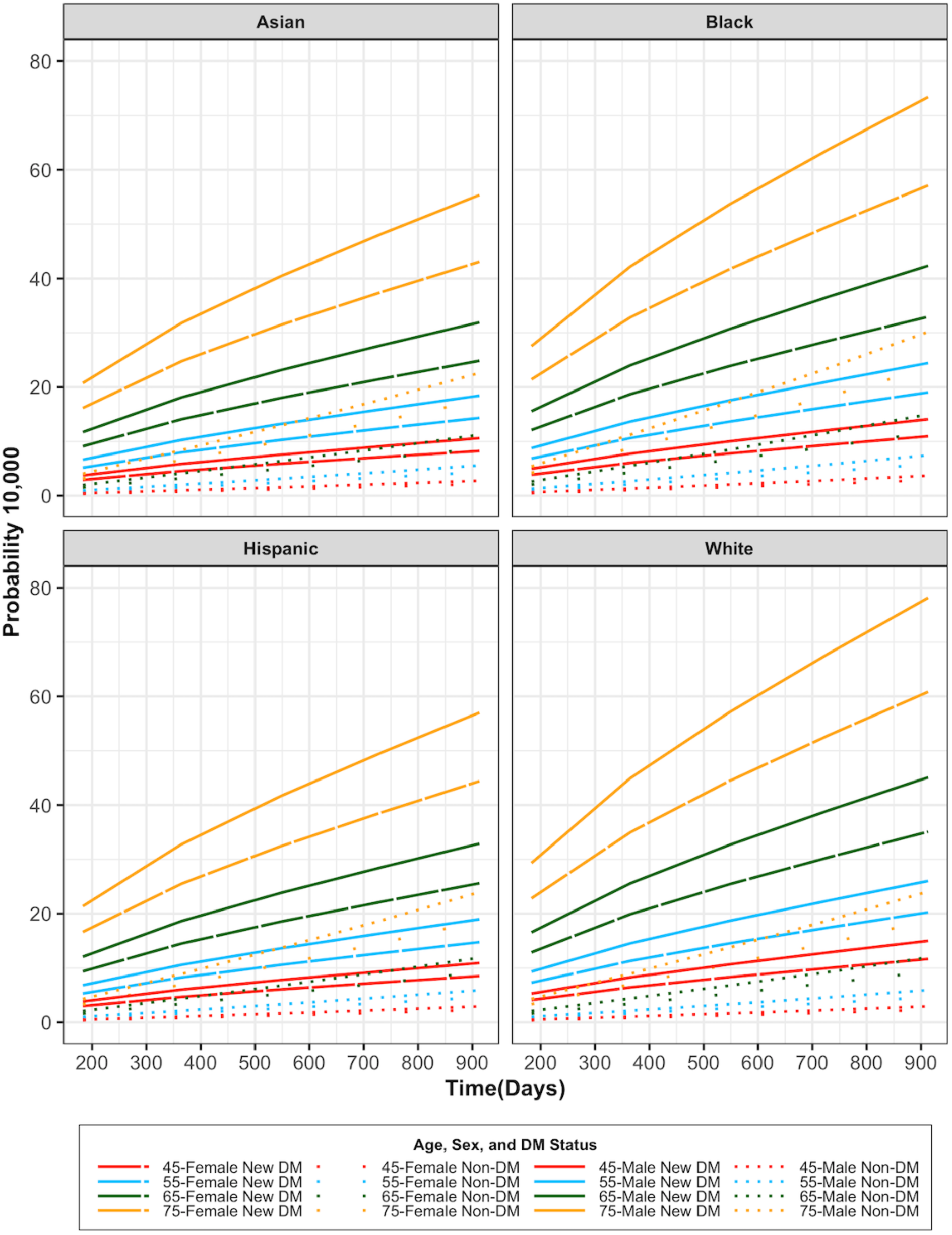
Cumulative probability of a pancreatic cancer diagnosis (%) by days since diabetes diagnosis/cohort entry by age, race, diabetes strata for selected ages (1-S_t_*100)

**Table 4:**
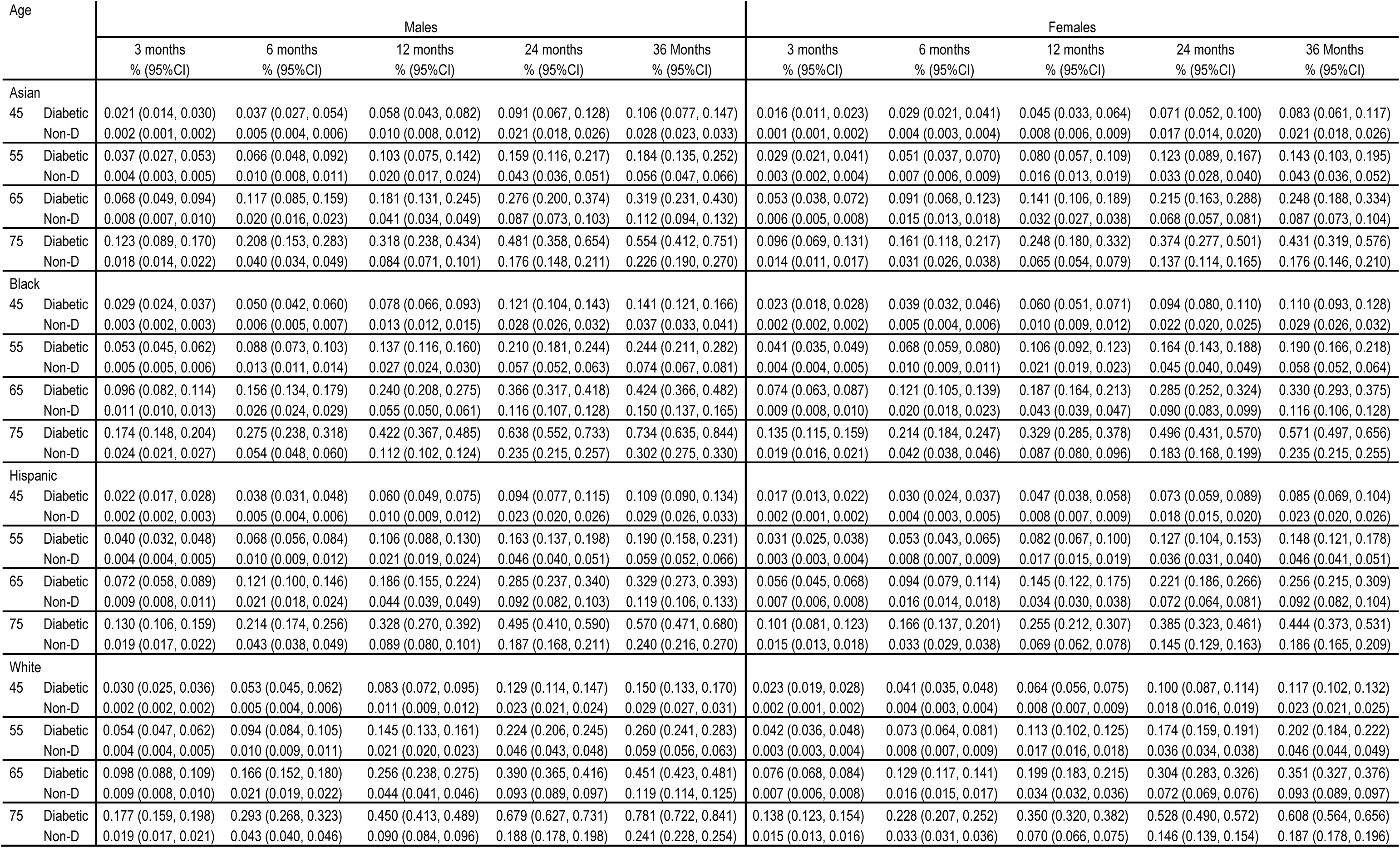
Cumulative probability (%) of pancreatic cancer by age, race, sex and time since diagnosis/cohort.

Individuals of Hispanic and Asian race who remained diabetes free had pancreatic cancer probabilities comparable to that of White individuals free of diabetes at the same timepoints, cumulative probability at one year of 0.089 (95%CI 0.080, 0.101) and 0.084 (95 5%CI 0.071, 0.101) for a 75-year old Hispanic and Asian male respectively. Yet Hispanic and Asian individuals with newly diagnosed diabetes were at a lower probability of pancreatic cancer than Black or White individuals with newly diagnosed diabetes, cumulative probability at one years of 0.328 (95%CI 0.270, 0.392) And 0.318 (95% CI 0.238, 0.434), respectively.

Because pancreatic cancer risk increases dramatically with age, cumulative probability of pancreatic cancer in older individuals without diabetes is comparable to the probability in younger individuals with a new diabetes diagnosis (Figure 2, Table 4), cumulative probability of pancreatic cancer for one year in a 45-year-old white who developed diabetes at age 45 was 0.083% (95%CI 0.072, 0.095). Based upon these probability estimates, we provide estimates of the number individuals with newly diagnosed diabetes who would need to undergo screening in order to detect one case of pancreatic cancer. These estimates assume a perfectly sensitive screening test (Stable3). The estimates of the number to be screened were slightly higher in Asian individuals and Hispanic individuals compared to Black individuals and White individuals. While the overall number needed-to-screen was lower in individuals with newly diagnosed diabetes compared individuals remaining diabetes free, this number varied considerably by age. For example, to detect one pancreatic cancer, one would have to screen 2,198 forty-five-year-old Asian females and 1,710 forty-five year-old Asian males at their time of diabetes diagnosis to detect one pancreatic cancer, assuming the screening test will only detect cancers that would present symptomatically within one year. This is about the same number of 75 year-old Asians without diabetes one would need to screen to detect one pancreatic cancer (1,527 and 1,188 in females and males, respectively). Similar patterns are seen for other racial groups (STable 3).

## DISCUSSION

Our goal was to estimate the age, race and sex-specific risk (Figure 2/Table 4) of pancreatic cancer among individuals with a new diabetes diagnosis and how hazard ratio (Figure 1/Table 2) decrease with time from diabetes diagnosis. Such estimates inform the window of opportunity and the potential number of pancreatic cancers that may be detected using a new-onset diabetes-based screening strategy. We did observe that the increased hazard ratio of pancreatic cancer was higher in younger individuals with newly diagnosed diabetics compared to individuals who developed diabetes at older ages. Furthermore, the hazard ratio also varied by race, with higher hazard ratios in White, Asian and Hispanic individuals compared to Black individuals. In the United States, the prevalence of all-cause diabetes also varies by age and race, with prevalence increasing with age and higher prevalence among Black individuals^28, 29^, and we observed these patterns in our data. In our analysis, the hazard ratio of pancreatic cancer in individuals newly diagnosed diabetics compared to individuals remaining free of diabetes followed the opposite pattern, such that the hazard ratio was lower in groups with a higher underlying prevalence of diabetes. This lower hazard ratio in groups at higher risk of diabetes could reflect a higher relative prevalence of diabetes from non-pancreatic cancer-related causes relative to pancreatic cancer-related diabetes. Like prior observational studies, we have shown a significantly higher rate of pancreatic cancer among individuals with newly diagnosed diabetes. However, our large sample size allowed us to closely examine how the hazard ratios of a pancreatic cancer diagnosis rapidly declined with time since the first diabetes diagnosis as compared to those remaining free of diabetes. For effective clinical screening, an understanding of the window of opportunity between the diagnosis of diabetes and when a diagnosis of pancreatic cancer based upon clinical symptoms would occur is critical to maximize benefit. In all groups, the hazard ratios of pancreatic cancer was greatest immediately following the initial diabetes claim and decreased rapidly over the following months, as shown in **Figure 1**. Thus, the maximum impact of screening is immediately following diabetes diagnosis. As the hazard ratio decreased with time, so too does the yield of screening, as most of the excess incidence occurs in the first few months. For example, the cumulative probability of pancreatic cancer two years after a diabetes diagnosis in Black men age 75 is 0.63% vs. 0.23% in non-diabetics, a difference of about 40 cancers per 10,000 individuals at 2 years; approximately 50% (n=21) of these “excess” cases occur in the first 6 months following the diabetes diagnosis and 75% (n=30) by the end of year one (Table 2). The true benefit of screening is further complicated by how quickly a newly-diagnosed diabetic individual could get screening for a potential pancreatic cancer, and to what extent detecting these cancers earlier would ultimately improve survival.

Our estimates of risk and hazard ratios include minority populations whereas most prior reported studies included predominantly white cohorts.^12, 15, 17, 30^ Setiawan et al conducted a large population-based cohort study of Black and Hispanic individuals , and found that a new-diabetes diagnosis was associated with a 3-4-fold increased risk of pancreatic cancer.^30^ Li et al examined diabetes and pancreatic cancer risk and included race and ethnicity, but their study was underpowered with respect to estimating interactions between age, sex and race.^17^ A large population-based cohort study from Kaiser Permanente Southern California with approximately 7.5 million person-years of follow-up found risk ratios for Hispanic and Asian individuals with a new diabetes diagnosis trended higher compared to Black and Whites individuals.^15^ In our analysis, we were well-powered to detect interactions by race and diabetes status. We found that non-diabetic Black have about a 25% higher risk of pancreatic cancer compared to White individuals, and there was little difference in risk between Asian, Hispanic and White individuals. However, among individuals with a new diabetes diagnosis, pancreatic cancer hazard ratio was about 30% higher in White individuals, compared with Black, Hispanic or Asian individuals (Table 2). It is important to note that the OLDW captures claims data from across the US among individuals with commercial insurance and Medicare Advantage. However, there is great regional diversity within racial groups in the US that may result in differences between our study and studies focused on a specific region. In addition, the results do not capture individuals without insurance, Medicaid, or Medicare (without Medicare Advantage)

Age-specific rates of pancreatic cancer have also been on the rise.^31^ Based on our study, the younger individuals with a new diabetes diagnosis had a higher hazard ratio compared to older individuals. However, the absolute risk of pancreatic cancer in younger newly-diagnosed diabetic individuals is very low because of their overall lower risk of pancreatic cancer. In contrast, in older individuals the hazard ratio of pancreatic cancer diagnosis following to a new diabetes diagnosis is lower, but because they have a much higher underlying risk of pancreatic cancer, their absolute risk is much higher. Absolute risk is a critical metric to consider with planning interventional screening studies.

Recent studies within the Nurse’s Health Study and Health Professionals Follow-up Study have demonstrated pancreatic cancer risk is highest in newly-diagnosed diabetic individuals who experience weight loss.^32^ A similar trend was observed in an analysis of Kaiser Permanente data which found more rapid increases in glucose levels, HbA1c levels and recent weight loss in patients with new diagnosis of diabetes who developed pancreatic cancer compared to those who did not.^15^ While EHR data is available within the Optum Labs Data Warehouse, it was available only for a fraction of the study population, limiting our ability to examine interactions by race and age.

Strengths of our study include the ability to examine interactions between race and diabetes status, as well as fully model the probability of pancreatic cancer as a function of time following diabetes diagnosis and how this probability varies by age. The Optum Labs Data Warehouse database includes broad representation of the United States population including a mixture of representation of ages. In addition, the use of Optum Labs Data Warehouse allows us to evaluate a relatively enrollees receiving care in different setting across widespread geographic areas of the United States.

There are several limitations to our study. First, our database relies on diagnosis based on ICD9, ICD10, and CPT-4 claims codes and as such we are not able to examine how factors that may underly diabetes such as obesity impact our probability estimates. Secondly, race/ethnicity was imputed using a proprietary algorithm. Prior validations of this imputation method have been previously published and demonstrated moderate sensitivity (48%), high specificity (97%) for Black race and high sensitivity (97%) and moderate specificity (47%) for whites^32^. If we applied these sensitivity and specificity estimates to our study population, the positive predictive value for race in the sample set is predicted to be 85% and 87% for Blacks and Whites respectively. Under the assumption that mis-classification of race is non-differential by diabetes status and pancreatic cancer, it is likely that this misclassification would attenuate our observed effects. Our model-based probabilities in non-diabetics are comparable to Surveillance, Epidemiology and End Results (SEER) rates (Stable 4)^33^ supporting the imputation algorithm. Our population included only those with commercial insurance and older individuals with Medicare Advantage, thus our results must be carefully considered when applying to individuals without insurance, Medicaid, or Medicare. Additionally, some individuals contributed to multiple enrollment periods, however our time-event model accounted for this and sensitivity analysis (randomly selecting 1 enrollment period per individual) yielded very similar results (results not shown). Future studies examining these findings in Medicare and Medicaid populations may be warranted.

In conclusion, in newly diagnosed diabetes, the probably of pancreatic cancer varied by age, race and sex, and time since diabetes diagnosis. While individuals with a new diabetes diagnosis do have a markedly increased risk of pancreatic cancer, the window of opportunity for earlier detection is short as the hazard rapidly with time following diabetes diagnosis, highlighting both the potential and challenges of early detection in this population.

## Data Availability

The data underlying the results of this study are third party data owned by Optum Labs and contain sensitive patient information; therefore the data is only available upon request. Interested researchers engaged in HIPAA compliant research may for data access requests. The data use requires researchers to pay for rights to use and access the data. All interested researchers can access the
data in the same manner as the authors - the authors had no special access privileges.

## Acknowledgements

This work was supported by the Stand Up To Cancer-Lustgarten Foundation Pancreatic Cancer Interception Translational Cancer Research Grant (Grant Number: SU2C-AACR-DT25–17).We would also like to acknowledge Nancy Porter for her assistance in this work.

