## Supplemental Materials for "Impact of race, sex and age on the probability of pancreatic cancer among patients with newly diagnosed diabetes"

Supplemental Table 1: Diagnosis/medications with equivalent codes

| Diagnosis | Codes |
|---|---|
| Diabetes | ICD-9 codes: 250.xx, 250.0x, 250.1x, 250.2x, 250.3x, 250.4x, 250.5x, 250.6x, 250.7x, 250.8x, 250.9x, 249.xx, V77.1*, 790.29*<br><br>ICD-10 codes: E10*, E10.1*, E10.2*, E10.3*, E10.4*, E10.5*, E10.6*, E10.8*, E10.9*, E11*, E11.1*; E11.2*; E11.3*, E11.4*, E11.5*; E11.6*, E11.8*; E11.9*, E08*, E08.0*, E08.1*, E08.2*, E08.3*, E08.36*, E08.37*, E08.39*, E08.4*, E08.5*, E08.6*, E08.8*, E08.9*, E13.0*, E13.1*, E13.2*, E13.3*, E13.36*, E13.37*, E13.39*, E13.4*, E13.5*, E13.6*, E13.8*, E13.9*, O24.4*, Z13.1*, R73.09. |
| Pancreatic Cancer | ICD-9 codes: 140.XX - 172.XX, 174.XX - 194.XX, 197.XX - 198.XX, 200.XX - 201.XX, 203.XX - 207.XX<br><br>(209.XX pancreatic neuroendocrine tumor)<br><br>ICD-10 codes: C00*-C43*, C45*-C75*, C77*-C79*, C81*-C94*, C96*-C99* (C25.4 pancreatic neuroendocrine tumor) |
| Pregnancy | ICD-9 codes 6310, 633, 6330, 63300, 63301, 6331, 63310, 63311, 6332, 63320, 63321, 6338, 63380, 63381, 6339, 63390, 63391, 640, 6408, 64080, 64081, 64083, 6409, 64090, 64091, 64092, 64093, 642, 6420, 64200, 6421, 64210, 6422, 64220, 6423, 64230, 64231, 64232, 64233, 64234, 6426, 64260, 64270, 6429, 64290, 643, 6432, 64320, 64321, 64233, 6438,64380, 64381, 64383, 6439, 64390, 64391, 64393, 645, 6450, 64510, 64511, 64513, 6452, ,64520, 64521, 64523, 646, 6461,64610, 64611, 64612, 6462, 64620, 64621, 64622, 6463, 64630, 64631, 64633, 6464, 64640,64641, 64642, 6465, 64650, 64651, 64652, 6466, 64660, 64661, 64662, 6467, 64671, 6468, 64680, 64681, 64682, 6469, 64690, 64691, 64693, 647, 64700, 64701, 64702, 6471, 64710, 6472, 64720, 6473, 64730, 6474, 64740, 6475, 64570, 6476, 64760, 6478, 64780, 6479, 64790, 648, 6480, 64800, 6481, 64810, 6482, 64820, 6483, 64830,6484, 64840, 6485, 64850, 6486, 6486, 6487, 64870, 6488, 64880, 6489, 64890, 649, 6490, 64901, 64902, 64903, 64904, 6491, 64910, 64911, 64912, 64913, 64914, 6492, 64920, 64921, 64922, 64923, 64924, 64953, 664930, 64931, 64932, 64933, 64934, 6494, 64940, 64941, 64942, 64943, 64944, 6495, 94950, 94951, 64953, 6410, 65100, 615101, 65103, 65104, 6511, 6511, 65111, 65113, 6512, 65120, 65121, 65123, 6513, 65130, 65131, 65133, 6514, 65140, 65141, 65143, 6515, 65150, 65151, 65153, 6516, 65161, 65163, 653, 6530, 65300, 6531, 65310, 65311, 65313, 6532, 65320, 65321, 65323, 6533, 65330, 65331, 65333, 654, 6540, 65410, 6542, 6545, 65450, 6546, 6546, 6547, 65470, 6548, 65480, 6549, 65490, 65500, 65510, 65520, 65530, 65540, 65560, 65610, 65620, 6581, 6581, 65811, 65813, 6594, 65940, 65943, 656050, 66400, 66410, 66420, 66430, 66440, 66450, 66480, 66490, 66520, 66530, 66543, 6655, 66560, 671, 6710, 67100, 6711, 67110, 6712, 67120, 6715, 67150, 6718, 67180, 6719, 67190, 67400, 67420, 677<br><br>ICD-10 Codes: O00 O000 O0000, O0001, O001, O0010, O00101, O00102, O00109, O001, O00111, O00112, O00119, O002, O0020, O00201, O00202, O00209, O0021, O00211, O00212, O00219, O008, O0081, O009, O0091, O0281, O04, O045, O046, O047, O048, O0480, O0481, O0482, O0483, O0484, O00485, O0486, O0487, O0488, O0489, O07, O070, O071, O072, O073, O0730, O0731, O0732, O0733, O0734, O0735, O0736, O0737, O0738, 0O0739, O074, O08, O081, O082, O083, O084, O085, O086, O087, O088, O0881, O0882, O0883, O0889, O089, O09, O090, O0900, O0901, O0902, O0903, O091, O0910, O0911, O0912, O0913, O092, O0921, O09212, O09213, O0919, O09229, O09219, O0929, O09291, O09292, O09293, O09299, O093, O0930, O0931, O0932, O0933, O094, O0940, O0941, O0942, O0943, O097, O0970, O0971, O0972, O0973, O0981, O09811, O09812, O09813, O09819, O0982, O09821, O09822, O09823, O09829, O0989, O09891, O09892, O09893, O09899, O0990, O0991, O0992, O0993, O09A0, O09A1 , O09A2, O09A3, O10, O100, O1001, O10011, O10012, O10013, O10019, O101, O1011, O10111, O10112, O10113, O10119, O102, O1021, O10211, O10212, O10213, O10219, O103, O1031. O10311, O10312, O10313, O10319, O104, O1041, O10411, O10412, O10413, O10419, O109, O1091, O10911, O10912, O10913, O10919, O12, O13, O131, O132, O133, O134, O135, O139, O150, O1500, O1502, O1503, O20, O208, O209, O21, O212, O218, O219, O22, O220, O2200, O2201, |

| | |
|---|---|
| | O2202, O2203, O221, O2210, O2211, O2212, O2213, O222, O2220, O2221, O2222, O2223, O223, O2230, O2231, O2232, O2233, O224, O2240, O2241, O2242, O2243, O225, O2250, O2251, O2252, O2253, O228, O228X, O228X1, O228X2, O228X3, O228X9, O229, O2290, O2291, O2292, O2293, O230, O2300, O2301, O2302, O2303, O231, O2310, O2311, O2312, O2313, O232, O2321, O2322, O2323, O233, O2331, O2332, O2333, O234, O2340 O2341, O2342, O2343, O235, O2351, O23512, O23513, O23519, O2352, O23521, O23522, O23523, O23529, O2359, O23591, O23592, O23593, O23599, O239, O2390, O2391, O2392, O2393, O24, O240, O2401, O24011, O24012, O24013, O24019, O241, O2411, O24111, O24112, O24113, O24119, O243, O2431, O24311, O24312, O24313 , O24319, O2441, O24410, O24414, O24415, O24419, O248, O2481, O24811, O24812, O24813, O24819, O249, O2491, O24911, O24912, O24913, O24919, O25, O251, O2510, O2511, O2512, O2513, O26, O260, O2600, O2601, O2602, O2603, O261, O261, O2611, O2612, O2613, O262, O2620, O2621, O2622, O2623, O263, O2630, O2631, O2632, O2633, O266, O2661, O26611, O26612, O26613, O26619, O267, O2671, O26711, O26712, O26713, O26719, O268, O2681, O26811, O26812, O26813, O26819, O2682, O26821, O26822, O26823, O26829, O2683, O26831, O26832, O26833, O26839, O2684, O2685, O26851, O26852, O26853, O26859, O2686, O2689, O26891, O26892 O26893, O269, O2690, O2691, O2692, O2693, O2699, O29, O290, O2901, O29011, O29012, O29013, O29019, O2902, O29021, O29022, O29023, O29029, O2909, O29091, O29092, O29093, O29099, O291, O2911, O29111, O29112, O29113, O29119, O2912, O29121, O29122, O29123, O29129, O2919, O29191, O29192, O29193, O29199, O292, O2921, O29211, O29212, O29213, O29219, O2929, O29291, O29292, O29293, O29299, O293, O293X, O293X1, O293X2, O293X3, O293X9, O294, O2940, O2941, O2942, O2943, O295, O295X, O295X1, O295X2, O295X3, O295X9, O296, O2960, O2961, O2962, O2963, O2969, O298, O298X, O298X1, O298X2, O298X3, O298X9, O299, O2991, O2992, O2993, O300, O3000, O30001, O30002, O30003, O30009, O3001, O30011, O30012, O30013, O30019, O3002, O30021, O30022, O30023, O30029, O3003, O30031, O30032, O30033, O30039, O3004, O30041, O30042, O30043, O30049, O3009, O30091, O30092, O30093, O30099, O301, O3010, O30101, O30102, O30103, O30109, O3011, O30111, O30112, O30113, O30119, O3012, O30121, O30122, O30123, O30129, O3013, O30131, O30132, O30133, O30139, O3019, O30191, O30192, O30193, O30199, O302, O3020, O30201, O30202, O30203, O30209, O3021, O30211, O30212, O30213, O30219, O3022, O30221, O30222, O30223, O30229, O3023, O30231, O30232, O30233, O30239, O3029, O30291, O30292, O30293, O30299, O311, O3110, O311X0, O311X1, O311X2, O311X3, O311X4, O311X5, O311X9, O311, O3111X, O3111X1, O3111X2, O3111X3, O3111X4, O3111X5, O3111X9, O3112, O3112X0, O3112X1, O3112X2, O3112X3, O3112X4, O3112X5, O3112X9, O3113, O3113X0, O3113X1, O3113X2, O3113X3, O3113X4, O3113X5, O3113X9, O312, O3120, O3120X0, O3120X1, O3120X2, O3120X3, O3120X4, O3120X5, O3120X9, O3121, O3121X0, O3121X1, O3121X2, O3121X3, O3121X4, O3121X5, O3121X9, O3122, O3122X0, O3122X1, O3122X2, O3122X3, O3122X4, O3122X5, O3122X9, O3123, O3123X0, O3123X1, O3123X2, O3123X3, O3123X4, O3123X5, O3123X9, O313, O3130, O3130X0, O3130X1, O3130X2, O3130X3, O3130X4, O3130X5, O3130X9, O3131, O3131X0, O3131X1, O3131X2, O3131X3, O3131X4, O3131X5, O3131X9, O3132, O3132X0, O3132X1, O3132X2, O3132X3, O3132X4, O3132X5, O3132X9, O3133, O3133X0, O3133X1, O3133X2, O3133X3, O3133X4, O3133X5, O3133X9, O367, O3670, O3670X0, O3670X1, O3670X2, O3670X3, O3670X4, O3670X5, O3670X9, O3671, O3671X0, O3671X1, O3671X2, O3671X3, O3671X4, O3671X5, O3671X9, O3672, O3672X0, O3672X1, O3672X2, O3672X3, O3672X4, O3672X5, O3672X9, O3673, O3673X0, O3673X1, O3673X2, O3673X3, O3673X4, O3673X5, O3673X9, O3680, O3680X0, O3680X1, O3680X2, O3680X3, O3680X4, O3680X5, O3680X9, O48, O480, O481, O8801, O88011, O88012, O88013, O88019, O8811, O88111, O88112, O88113, O88119, O8821, O88211, O88212, O88213, O88219, O8831, O88311, O88312, O88313, O88319, O8881, O88811, O88812, O88813, O88819, O91, O910, O9101, O91011, O91012, O91013, O9019, O911, O9111, O91111, O91112, O91113, O91119, O92, O920, O9201, O92011, O92012, O92013, O92019, O921, O9211, O92111, O92112, O92113, O92119, O922, O9220, O9229, O94, O98, O980, O9801, O98011, O98012, O98013, O98019, O981, O9811, O98112, O98113, O98119, O982, O9821, O98211, O98212, O98213, O98219, O983, O9831, O98311, O98312, O98313, O98319, O984, O9841, O98411, O98412, O98413, O98419, O985, O9851, O98511, O98512, O98513, O98519, O986 O9861, O98611, O98612, O98613, O98619, O987, O9871, O98711, O98712, O98713, O98719, O988, O9881, O98811, O98812, O98813, O98819, O989, O9891, O98911, O98912, O98913, O98919, O99, O990, O9901, O99011, O99012, O99013, O99019, O991, O9911, O99111, O99112, O99113, O99119, O992, O9921, O99210, O9911, O99212, O99213, O9928, O99280, O99281, O99282, O99283, O993, O9931, O99310, O99311, O99312, O99313, O9932, O99320, O99321, O99322, O99323, O9933, O9930, O9933, O99332, O99333, O9934, O99340, O99341, O99342, O99343, O9935, O99350, O99351, O99352, O99353, O994, O9941, O99411, O99413, O99419, O995, O9951, O99511, O99512, O99513, O99518, O996, O9961, |

| | |
|---|---|
| | O99611, O99612, O99613, O99619, O997, O9971, O99711, O99712, O99713, O99719, O998, O9981, O99810, O9982, O9983, O99830, O9984, O99840, O99841, O99842, O99843, O9989, O9A, O9A1, O9A11, O9A111, O9A112, O9A113, O9A119, O9A2, O9A21, O9A211 O9A212, O9A213, O9A219, O9A3, O9A31, O9A311, O9A312, O9A313, O9A319, O9A4, O9A41, O9A411, O9A412 O9A413 O9A419, O9A5, O9A51, O9A511, O9A512, O9A513, O9A519, V1521, V22, V220, V221, V23, V23, V230, V231, V232, V233, V234, V2341, V2342, V2349, V234, V238, V2381, V2382, V2383, V2384, V2385, V2386, V2387, V2389. V239, V616, V617, V7242, Z3201, Z332 Z34, Z340, Z3400, Z3401, Z3402, Z3403, Z348, Z3480, Z3481, Z3482, Z3483, Z349, Z3490, Z3491, Z3492, Z3493, Z3A0, Z3A00, Z3A01, Z3A08, Z3A09, Z3A1, Z3A10, Z3A11, Z3A12, Z3A13, Z3A14, Z3A15, Z3A16, Z3A17, Z3A18, Z3A19, Z3A2, Z3A20, Z3A21, Z3A22, Z3A23, Z3A24, Z3A25, Z3A26, Z3A27, Z3A28, Z3A29, Z3A3, Z3A30, Z3A31, Z3A32, Z3A33, Z3A34, Z3A35, Z3A36, Z3A37, Z3A38, Z3A39, Z3A4, Z3A40, Z3A41, Z3A42, Z3A49, Z640. |
| Insulin | CPT codes: A4224, A9274, G9147, J1815, J1817, J1820, K0548, S5550, S5551, S5552, S5553, S5560, S5561, S5565, S5566, S5570, S5571, S8490, S9353, A4225, A4230, A4231, A4232, E0784, S9145<br><br>ICD-9 codes: V58.67, V45.85, V53.91, V65.46, 996.57, 99.17<br><br>ICD-10 codes: Z79.4, T85.614*, T85.624*, T85.633*, T85.694*, 3E013VG, 3E030VG, 3E033VG, 3E040VG, 3E043VG, 3E050VG, 3E053VG, 3E060VG, 3E063VG |
| Diagnosis (including Pathology, Radiology and Laboratory Procedures) | CPT codes: 36400-36425, 70010-76999, 78000-78799, 80000-89999<br><br>HCPCS: S9529, G0001<br><br>AMA Site Code: 81 |

Supplemental Table 2: Study population inclusion and exclusions.

Diabetes Cohort:

| Attrition Steps | Individuals remaining |
|---|---|
| Individuals identified with at least 1 claim for diabetes, screening, abnormal glucose, OAD or insulin between 2008 and Sept2018 | 6,503,075 |
| Individuals with at least 2 claims (at least one of which is a diabetes diagnosis) | 3,513,882 |
| Individuals with 1 year continuous enrollment and no baseline evidence for diabetes, screening, abnormal glucose, OAD or insulin | 527,435 |
| Individuals at least 40 years old | 460,033 |
| Individuals with known demographic information (business line and gender) | 459,604 |
| Cancer exclusions and Pregnancy exclusion | 424,129 |

**Non-exposed diabetes Cohort:**

| Attrition Steps | Enrollment periods remaining |
|---|---|
| Enrolled (unique CE periods) between 2008 and Sept2018 | 23,270,021 |
| Unique CE periods without diabetes in 1 year prior to "derived" index date | 12,659,694 |
| Unique CE periods where individual is least 40 years old | 5,723,768 |
| Unique CE periods where individual has known demographic information (business line and gender) | 5,721,956 |
| Unique CE periods where individual meets cancer exclusions and pregnancy exclusion | 5,420,805 |

Supplemental Table 3: Number needed to screen to detect one pancreatic cancer case assuming perfect screening test for cancers that would present symptomatically in 1 year by age, race, sex and diabetes status.

| | **Newly Diagnosed Diabetics Males/Females** | | **Males/Females without diabetes** | |
|---|---|---|---|---|
| | Males | Females | Males | Females |
| Asian | | | | |
| Age 45 | 1,710 | 2,198 | 10,140 | 13,037 |
| Age 55 | 972 | 1,249 | 4,961 | 6,378 |
| Age 65 | 552 | 710 | 2,427 | 3,120 |
| Age 75 | 314 | 404 | 1,188 | 1,527 |
| Black | | | | |
| Age 45 | 1,289 | 1,657 | 7,594 | 9,764 |
| Age 55 | 732 | 942 | 3,716 | 4,777 |
| Age 65 | 416 | 535 | 1,818 | 2,337 |
| Age 75 | 237 | 304 | 890 | 1,144 |
| Hispanic | | | | |
| Age 45 | 1,660 | 2,134 | 9,564 | 12,297 |
| Age 55 | 943 | 1,213 | 4,679 | 6,016 |
| Age 65 | 536 | 689 | 2,289 | 2,943 |
| Age 75 | 305 | 392 | 1,120 | 1,440 |
| White | | | | |
| Age 45 | 1,210 | 1,556 | 9,520 | 12,240 |
| Age 55 | 688 | 884 | 4,657 | 5,988 |
| Age 65 | 391 | 503 | 2,279 | 2,930 |
| Age 75 | 222 | 286 | 1,115 | 1,424 |

Supplemental Table 4: SEER 1-year Incidence Rates for Pancreatic Cancer (%) from 2008-2018 and 1-year estimated Probability of pancreatic cancer in non-diabetics.

| | Male | | Female | |
|---|---|---|---|---|
|  | **SEER** % | **OLDW enrollees without diabetes % (95%CI)** | **SEER** % | **OLWD enrollees without Diabetes % (95%CI)** |
| **Asian** | | | | |
| 45 | 0.004 | 0.010 (0.008, 0.012) | 0.002 | 0.008 (0.006, 0.009) |
| 55 | 0.012 | 0.020 (0.017, 0.024) | 0.009 | 0.016 (0.013, 0.019) |
| 65 | 0.036 | 0.041 (0.034, 0.049) | 0.027 | 0.032 (0.027, 0.038) |
| 75 | 0.061 | 0.084 (0.071, 0.101) | 0.061 | 0.065 (0.054, 0.079) |
| **Black** | | | | |
| 45 | 0.007 | 0.013 (0.012, 0.015) | 0.006 | 0.010 (0.009, 0.012) |
| 55 | 0.029 | 0.027 (0.024, 0.030) | 0.020 | 0.021 (0.019, 0.023) |
| 65 | 0.070 | 0.055 (0.050, 0.061) | 0.053 | 0.043 (0.039, 0.047) |
| 75 | 0.105 | 0.112 (0.102, 0.124) | 0.076 | 0.087 (0.080, 0.096) |
| **Hispanic** | | | | |
| 45 | 0.004 | 0.010 (0.009, 0.012) | 0.003 | 0.008 (0.007, 0.009) |
| 55 | 0.015 | 0.021 (0.019, 0.024) | 0.012 | 0.008 (0.007, 0.009) |
| 65 | 0.039 | 0.044 (0.039, 0.049) | 0.032 | 0.034 (0.030, 0.038) |
| 75 | 0.071 | 0.089 (0.080, 0.101) | 0.066 | 0.069 (0.062, 0.078) |
| **White** | | | | |
| 45 | 0.005 | 0.011 (0.009, 0.012) | 0.003 | 0.008 (0.007, 0.009) |
| 55 | 0.019 | 0.021 (0.020, 0.023) | 0.014 | 0.017 (0.016, 0.018) |
| 65 | 0.050 | 0.044 (0.041, 0.046) | 0.032 | 0.034 (0.032, 0.036) |
| 75 | 0.084 | 0.090 (0.084, 0.096) | 0.066 | 0.070 (0.066, 0.075) |
